# Published registered reports are rare, limited to one journal group and inadequate for randomized controlled trials in the clinical field

**DOI:** 10.1101/2022.08.02.22278318

**Authors:** Norah Anthony, Antoine Tisseaux, Florian Naudet

## Abstract

**Objective:** Registered reports relate to a new publication of a peer-review of the protocol before the start of the study, followed by an in-principle acceptance by the journal before the study starts. We aimed to describe randomized controlled trials (RCTs) in the clinical field published as registered reports.

**Study design and setting:** This cross-sectional study (registration: https://osf.io/zf53p/) included registered report results for RCTs, identified on PubMed/Medline and on a list compiled by the Center for Open Science. It explored the proportion of reports that received in-principle acceptance (and/or published a protocol before inclusion of the first patient) and changes in the primary outcome.

**Results:** A total of 93 RCT publications identified as registered reports were included. All but one were published in the same journal group. The date of the in-principle acceptance was never documented. For most of these reports (79/93, 84.9 %) a protocol was published after the date of inclusion of the first patient. A change in the primary outcome was noted in 40/93 (44%) of these publications. Three out of the 40 (33%) mentioned this change.

**Conclusions:** Randomized controlled trials in the clinical field identified as registered reports were rare, they originated from a single journal group and did not comply with the basic features of this format.

**Protocol registration:** https://osf.io/zf53p/

**What is new ?:**

- The registered report format for clinical randomized controlled (RCTs) trials is still marginal and few journals make use of it.
- The clinical RCTs identified as registered reports were from a single journal group and did not necessarily comply with the basic features of this format, and common biases may thus persist.
- To improve research trustworthiness, more efforts need to be made by Journal publishers, trial funders, etc. for the implementation of this format for clinical RCTs.

## Introduction

Clinical trial results shape clinical practice guidelines and support decisions that affect the health of millions of people. According to clinicaltrials.gov, “in a clinical trial, participants receive specific interventions according to the research plan or protocol created by the investigators. These interventions may be medical products, such as drugs or devices; procedures; or changes to participants’ behavior, such as diet.” Randomization is a method used to randomly assign participants to the arms of a clinical study and is a key to ensuring the initial comparability of the groups. Following the Thalidomide crisis in 1962 (1), randomization became a gold standard for clinical trials, in order to maximize their internal validity. Because their results must be trustworthy and resist the considerable financial and ideological conflicts of interest inherent in the evaluation of therapeutics, clinical randomized controlled trials (RCTs) have long been at the forefront of efforts to ensure transparency and reproducibility. In 2005, to enhance transparency, the International Committee of Medical Journal Editors (ICMJE) made the registration of clinical RCTs compulsory before inclusion of the first patient (2). However, and despite these efforts, many issues remain. Many clinical RCT results remain unpublished, especially when conclusions are “negative” (3). When published, clinical RCT results are frequently reported with a certain degree of selective reporting and spin (4). Initiatives promoting clinical RCT data-sharing are too few (5). In addition, clinical RCTs all too often ask the wrong question, e.g. by relying on the wrong comparator, and/or they implement inadequate methods, e.g. the use of non-informative surrogate outcomes, or they lack adequate power (6). All these problems hamper reproducibility and reduce the value of therapeutic research. Similar concerns have been described in many scientific disciplines, such as psychology (7) and cancer biology (8), suggesting the existence of a reproducibility crisis in science (9).

A new publication format – registered reports – was created to address these systemic flaws by increasing transparency and relying on a strict hypothetical-deductive model of scientific method (10,11). In a registered report, the study protocol – including theory, hypotheses, detailed methods, and analysis plan – is submitted directly to a journal, and peer review of this material – i.e. stage-1 peer review – occurs before the research is underway. Stage-1 peer review appraises both the importance of the research question and the quality of the methodology prior to data collection (12–14). Following stage-1 peer review, high quality protocols exploring relevant research questions receive an in-principle acceptance for publication in the journal, on condition that the study is completed successfully in accordance with the protocol. Provisional acceptance is therefore given without knowledge of the research results, or whether or not the study hypothesis has been verified. Following an in-principle acceptance, the investigators can register their protocol and start the research. Once the research is completed, stage-2 peer review checks its compliance with the protocol and the accuracy of the conclusions in view of the evidence. Figure 1 (adapted from the figure provided by the Center For Open Science (12)) gives details of the whole registered report process. This approach is expected to promote high methodological quality and to reduce issues such as publication bias. In addition stage-1 peer reviews, if qualitative, can deal with certain issues regarding design pre-emptively, and this should improve the robustness of the methods. Stage-2peer review ensures compliance with the methodology validated in stage 1 and leads to the publication of the study whatever the results. This approach is expected to limit publication bias and selective outcome reporting. Registered reports also have the advantage of pre-emptively defining the terms of important issues such as data sharing, which should indeed be addressed before the first patient is included in order to make sure that trial participants are adequately informed. This process is therefore quite different from the publication of a protocol. Some journals (e.g. BMJ Open, Plos One, Trials, etc.) offer the opportunity to publish protocol papers for ongoing trials, which provide more details than clinical trial registries. However, while publishing the protocol is generally considered as good practice in terms of research transparency, it does not completely remove the possibility of publication bias or selective outcome reporting bias in the final report. In addition, unlike the registered report format, the publication of a protocol often occurs after the study has started and this does not allow any issues to be resolved regarding the research question and design on the basis of the reviewers’ comments. Furthermore, in some cases, there is no specific peer review by the journal for these protocols, for instance at BMJ Open, acceptance is offered providing the study 1/ was funded by a funding body ensuring peer review and 2/ obtained authorization from an appropriate Institutional review board (15). Finally, it is also worth noting that the use of registered reports in journals is generally associated with other open science practices (16).

**Figure 1.**
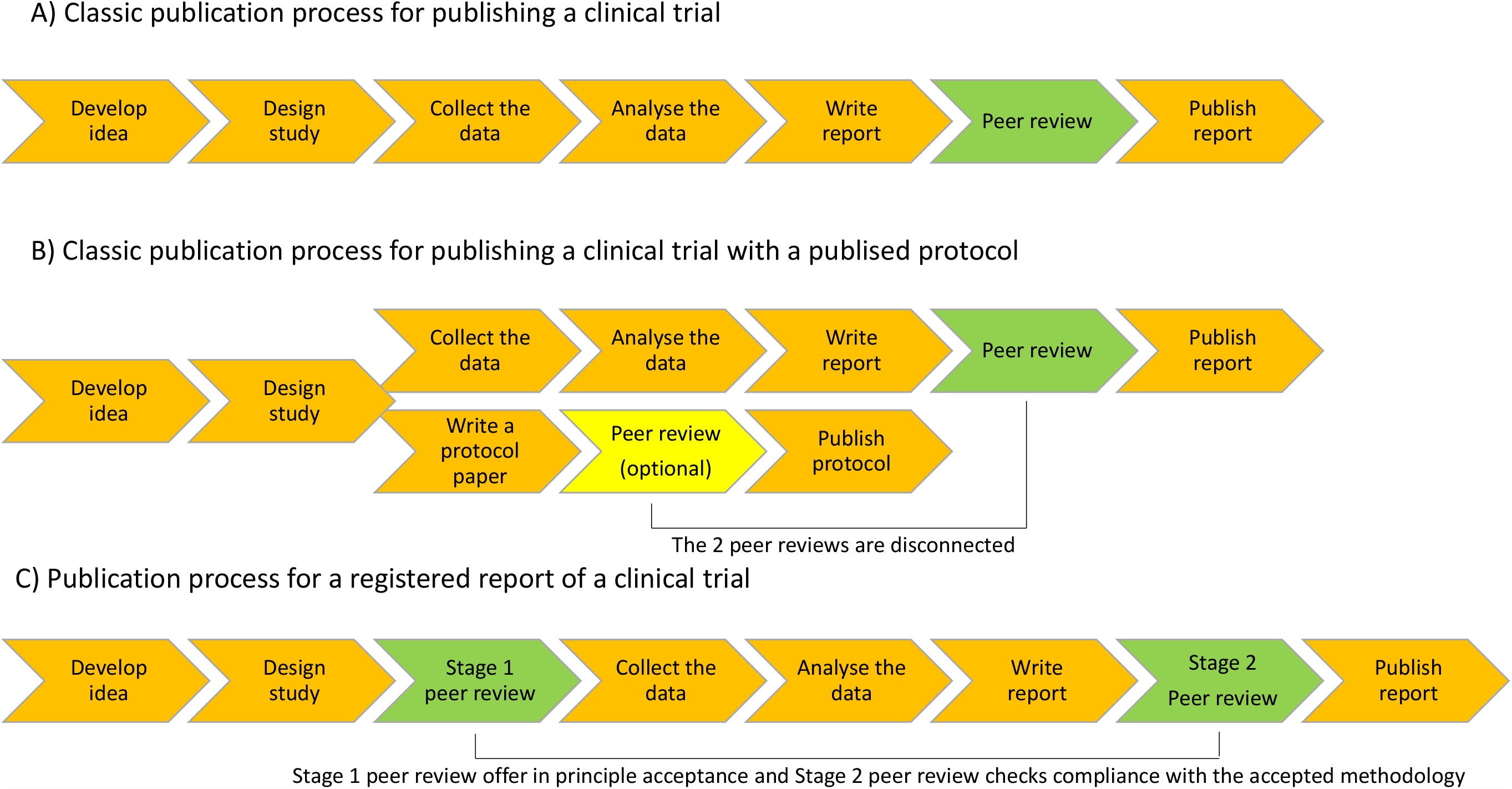
Chart representing different publication processes for a clinical trial and its protocol

To date, more than 300 scientific journals, mainly, but not only, from the field of psychology (11), have adopted the registered report format. These journals include PLOS Biology, BMJ Open Science, Nature and Nature Human Behavior. Despite the adequate fit between the requirements of the registered format and the confirmatory nature of clinical RCTs (14), few medical journals have implemented registered reports. BMC Medicine launched the first registered report format for RCTs in 2017 (14). The mega-journal PLOS ONE, which occasionally publishes RCTs, has adopted the registered report format(17). The specialist journals affiliated to the Journal of Medical Internet Research (JMIR) also accept registered reports and assign them an International Registered Report Identifier (IRRID) number (18). Our objective was to identify and describe features of clinical randomized controlled trials published as Registered Reports.

## Method

The methods of this cross-sectional study were specified in advance. They were documented in a protocol registered on the Open Science Framework on 17^th^ December 2021 (https://osf.io/zf53p/).

### Eligibility criteria

We surveyed clinical RCTs, i.e. randomized controlled trials that had a clinical orientation, published as final reports of a registered report. Registered report protocols (i.e. stage-1 registered reports) were not included.

### Search strategy and study selection

The search for these registered reports used the Medline database with the following search string: ““registered report” or “IRRID”. Following peer review, we also performed a sensitivity analysis using the terms “registered replication report*” and “preregistered report*. In addition, we searched a list of published registered reports in a Zotero library compiled by the Center for Open Science https://bit.ly/2pJRYz3. Database searches were performed on 17th December 2021.

All references identified were first automatically screened using R, excluding references where the title included the word “protocol” and/or did not include the character string “random”. This procedure was validated on 100 randomly sampled references, retrieving 100 % accuracy before being implemented (details of the validation procedure are shown in the **Web Appendix 1**). All remaining references were then manually selected by two independent reviewers (NA and AT). Any disagreement was resolved by consensus with FN.

### Outcomes

#### Primary outcome

The primary outcome was the proportion of RCTs identified as registered reports that had received in-principle acceptance and/or had published a protocol before inclusion of the first patient.

Additional information was collected on i) the protocol (protocol availability, date of the time-stamped protocol, date of protocol submission, date of protocol acceptance, date of protocol publication in a journal and publication of the protocol in the same journal or journal group), ii) the primary outcome used in the registered report (change in the primary outcome, and in case of a change, mention of the change in the registered report), iii) the secondary outcomes used in the registered report (change in the secondary outcomes and in case of a change, mention of the change in the registered report), iv) result from the study for its primary outcome (positive or negative), v) sample size, vi) power to detect effect sizes of 0.3, 0.5 and 0.8, respectively (based on sample size in each arm of the trial), vii) citation rate, viii) Altmetric attention score and ix) characteristics of the registered report (journal, journal impact factor and topic).

### Data extraction

Two authors (NA and AT) independently extracted the data from the studies included clinical trial registry data and PubMed metadata on the articles (see **Web Appendix 2**). Disagreements were resolved by consensus or in consultation with a third reviewer (FN). Data extraction for the number of citations and the Altmetric attention score was performed on the 27th April 2022.

### Statistical methods

A descriptive analysis was performed using the following: medians (and range) for quantitative variables, numbers and percentages for qualitative variables. Statistical analyses were performed using R version 4.0.3.

### Deviations from the protocol or addition of elements

For the sake of simplicity, the power calculation was limited to parallel RCTs, which accounted for most of the registered reports. Since the date of the first inclusion of patients was only documented as month and year, we computed time lapses using months as the unit.

Because we identified several publications during our searches which, despite being identified as registered reports, were secondary analyses and not the report on the primary analysis of the trial (e.g. based on the pre-specified analysis of its primary outcome), we added this feature (primary/secondary analysis) as one of our outcomes and provided detailed results according to this variable.

### Patient involvement

We had no established contacts with specific patient groups who might be involved in this project. No patients were involved in defining the research question or the outcome measures, nor were they involved in the design and implementation of the study. There are no plans to involve patients in the dissemination of the results, nor will we disseminate the results directly to patients.

## Results

A total of 2074 references were identified from Zotero (n=194) and Medline (n=1880). Regarding the sensitivity analysis, no additional paper matched the search criteria. One hundred and fifteen references remained after deduplication and automatic screening. Of these, 22 were not identified as registered reports, and 93 references were eligible for inclusion because they were identified as registered reports and/or had an IRRID (**Figure 2**). Fifty-three out of 93 (57%) of the reports were published in the JMIR and an additional 39 (42 %) were published in a JMIR-affiliated journal, leaving only one other report, published in Nurse Educ Today. Of the 93, 78 (83.9 %) reported results on the primary trial analysis and 15 (16.1 %) reported on a secondary analysis.

**Figure 2.**
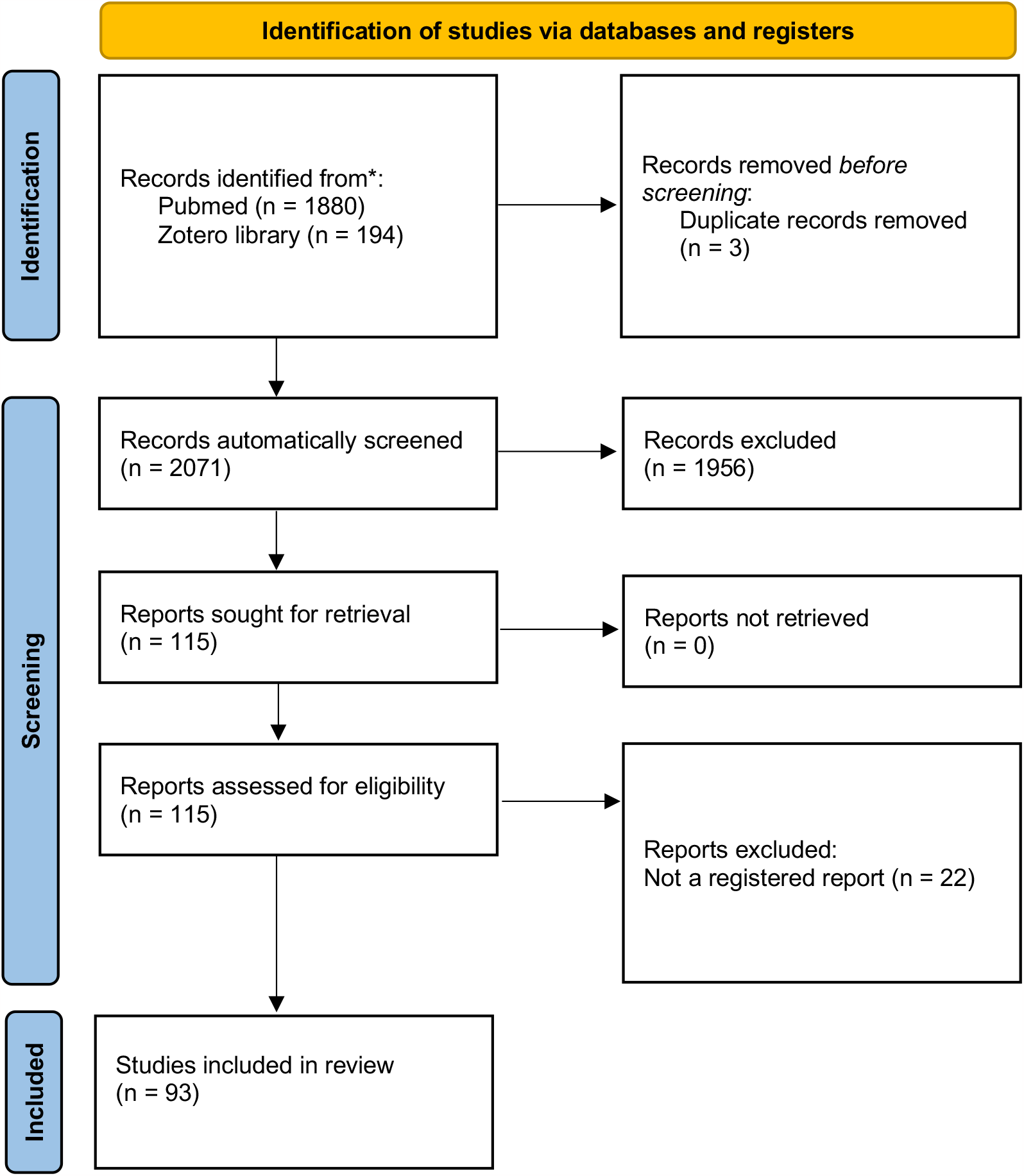
Flow chart for registered report selection

### Primary outcome

The date of the in-principle acceptance was never documented in the reports included. Consequently, we relied on the date of protocol publication. We identified protocols published in a journal before inclusion of the first patient for 8/93 reports and protocol acceptance before the first inclusion for 8/93 reports. The lack of dates prevented any checks regarding protocol publication and protocol acceptance for an additional 6/93 and 14/93 registered reports respectively. In other words, 79/93 (84.9 %) had a protocol published after the date of inclusion of the first patient and 71/93 (76.3%) had a protocol accepted after the first inclusion. **Figure 3** reports the results for our primary outcome in the sample of 78/93 (83.9%) registered reports reporting results from primary analyses. In this sample, 7/78 (9%) had a protocol published in a journal before inclusion of the first patient, 5/78 (6.4 %) had missing data for this item and 66/78 (84.6 %) had a protocol published after the date of inclusion of the first patient. Among the registered reports on primary analyses, protocol publications ranged from 1.4 years before to 3 years after the inclusion of the first patient (the median time lapse was 1 year); of these 78, 42 (53.8%) had their protocols published up to one year after the first inclusion. Fifty-two out of 78 (66.7%) were registered in a trial registry before inclusion of the first patient, 21/78 (26.9 %) were registered after inclusion of the first patient, and dates were not available for 5/78 (6.4 %).

**Figure 3.**
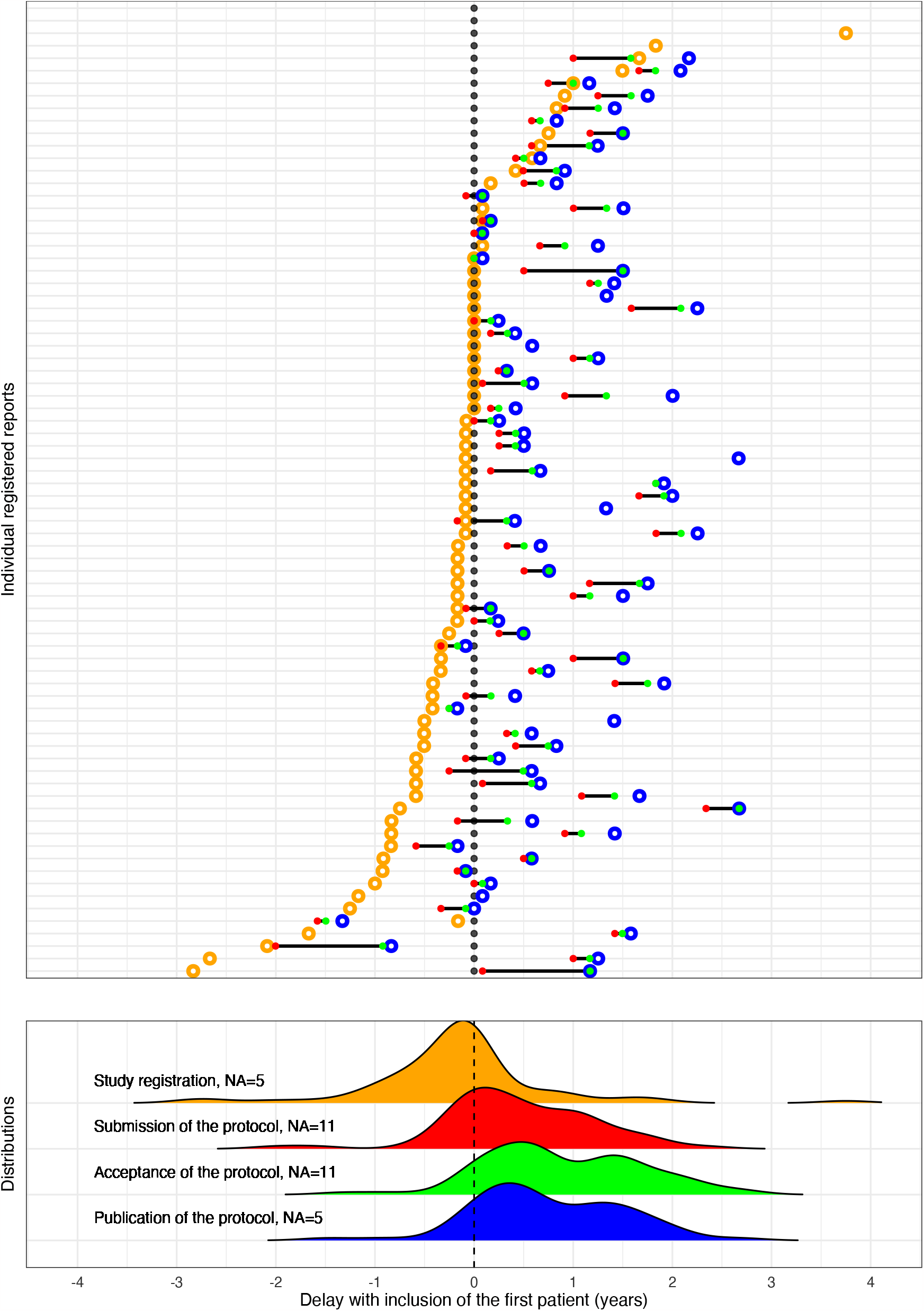
Chart representing clinical randomized controlled trial registration (*orange*), submission of the protocol (*red*), acceptance of the protocol (*green*), and protocol publication (*blue*) in relation to the date of inclusion of the first patient (*black*)

### Secondary outcomes

A protocol was available for 90/93 (97%) reports and all of these 90 protocols were published in a journal. The published protocols and their associated reports were not published in the same journal group for 55 (61%) of these 90. We found no mention of OSF registration in any of these papers. A change in primary outcome compared to the protocol was noted for 40/93 (44%) of the reports, including 27/78 (35%) of the reports reporting the results from primary analyses. Among these 40 registered reports, we identified 9 that added components to the primary outcomes, 7 that deleted components of the outcomes, 1 that switched the primary outcome with a secondary outcome and 23 that modified the definition of the primary outcome (7, 3 and 13 respectively for a change of period of assessment, complete change of criteria and secondary analysis of RCT data). Thirteen out of 40 (33%) of the reports with a change in the primary outcome mentioned this change, and these 13 reports were exclusively secondary analyses.

A change in at least one of the secondary outcomes was present in 46/87 (53%) reports of studies with secondary outcomes (absence of secondary outcomes in 5 protocols/reports) and this change was mentioned by 8/46 (17%) of them.

**Table 2** provides details for these results with a specific focus on reports providing results from primary analyses versus those only reporting secondary analyses. The study outcomes were considered positive, mixed and negative in 49 (52.7%), 10 (10.8%) and 34 (36.6%) of the reports included respectively. The median sample size was 239 (range 13-14 482) for the 93 reports. Over the 75 trials with a parallel design, the median power (interquartile range) was 51% (26%-86%), 91% (59%-99%), 99% (94%-100%) to detect effect sizes of 0.3, 0.5 and 0.8, respectively (**Figure 4**). The median Altmetric score was 2 (0-574) and the median citation rate was 2 (0-26), for a median of 374 (206-528) days after publication. Descriptive statistics for all other outcomes are shown in **Table 1**.

**Table 1.**
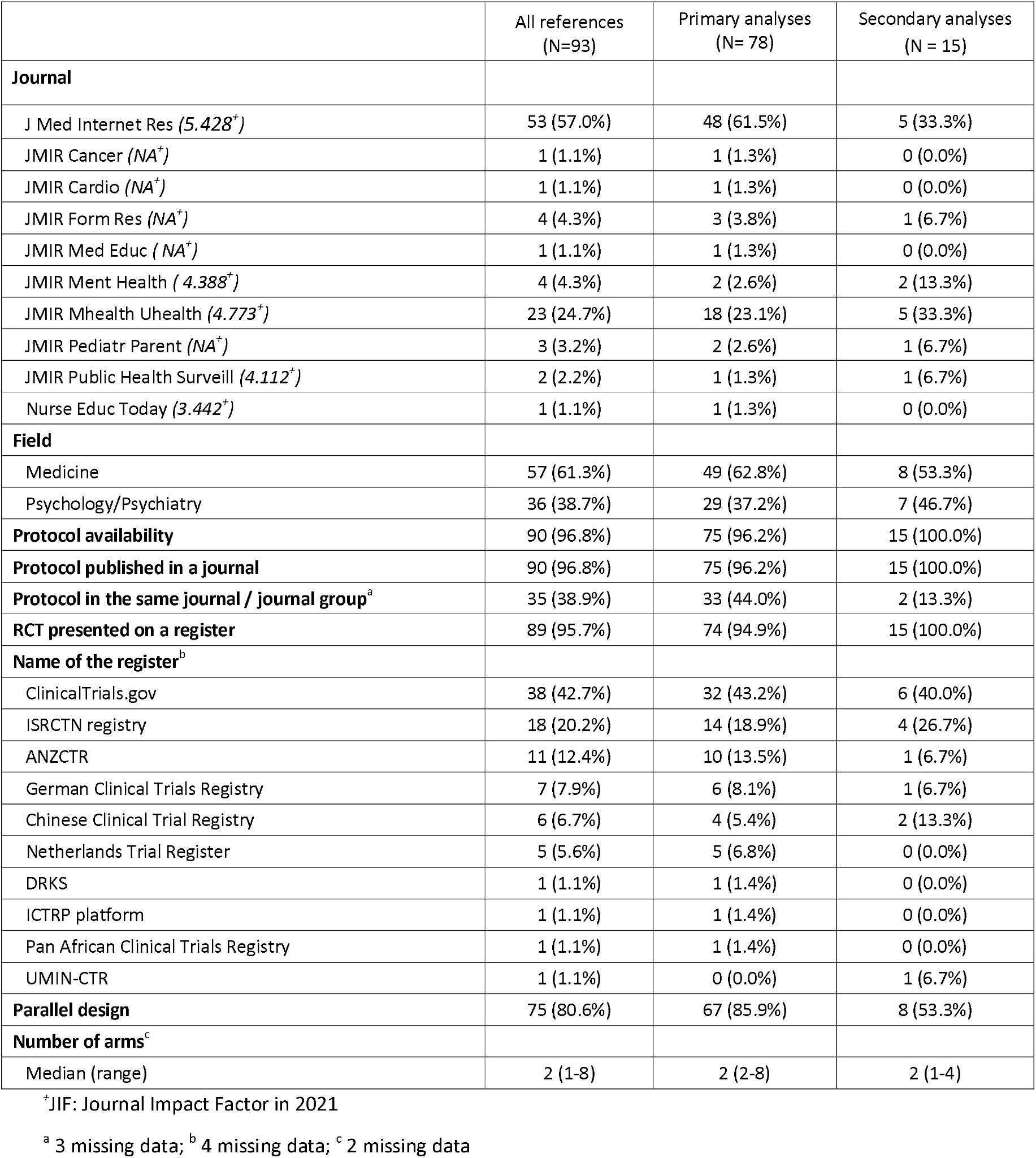

**Table 2.**
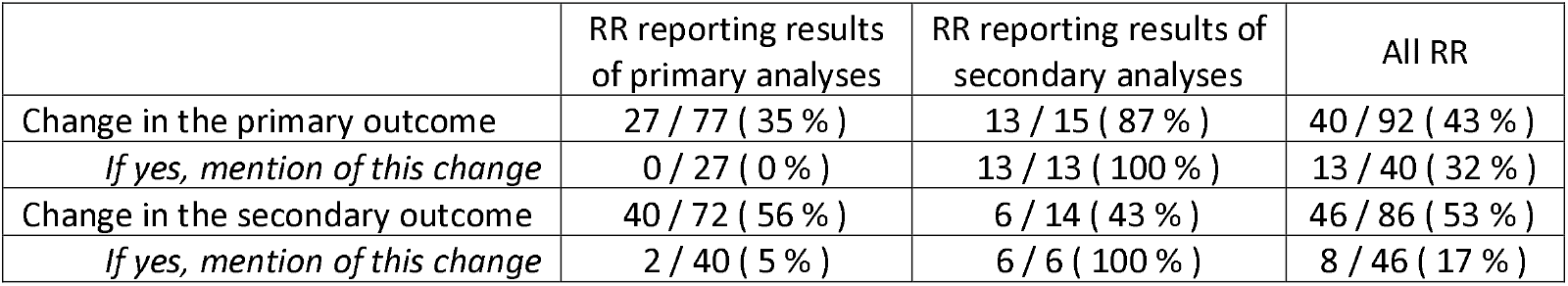

**Figure 4.**
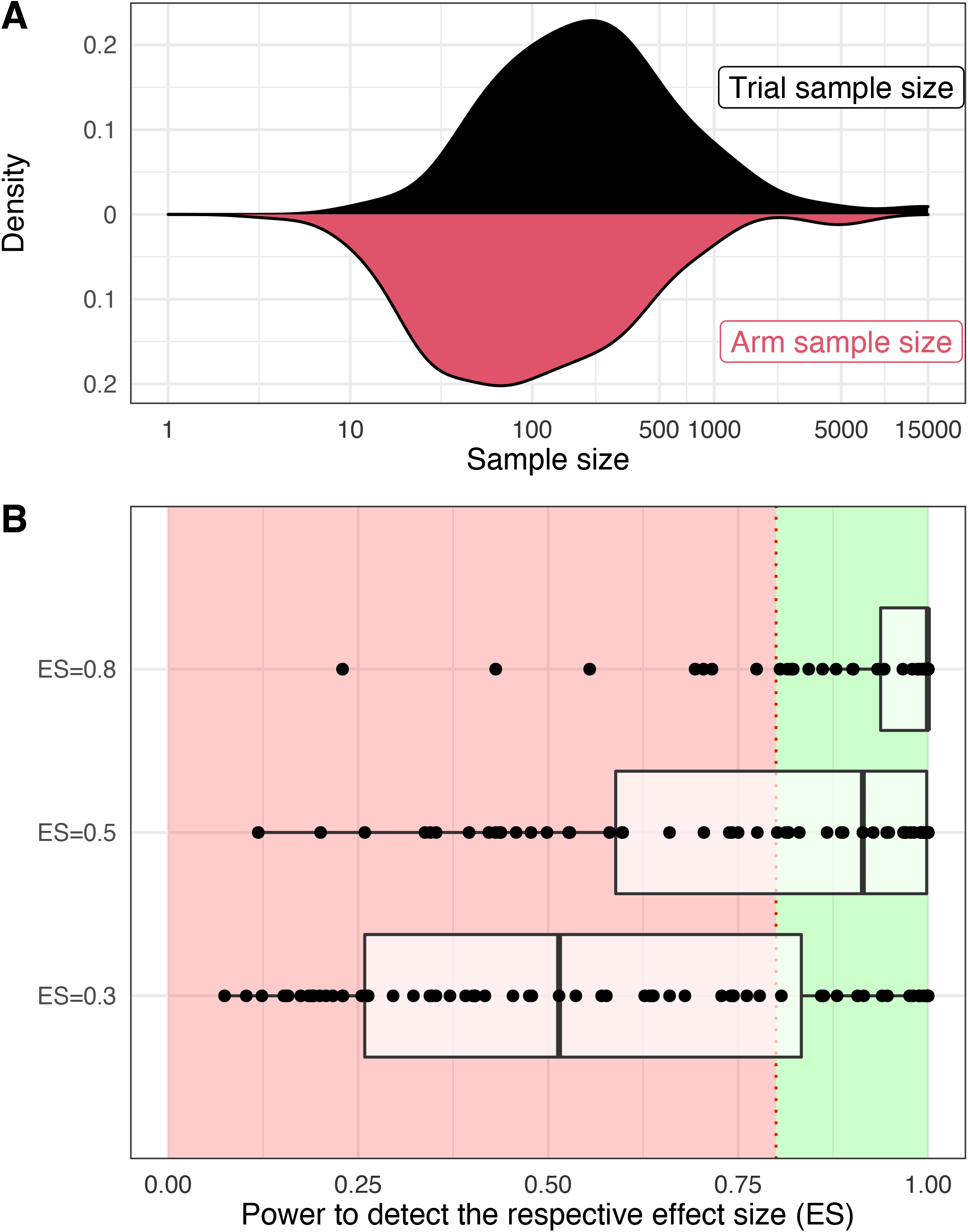
Power analyses A. Density curves representing trial sample size distribution and arm sample size distribution B. Box plot representing power values to detect three different effect sizes

## Discussion

### Statement of principal findings

In short, the registered report format is virtually non-existent for published RCTs, except in the JMIR group. To date, BMC Medicine has only published 3 registered reports (19–21), and none concerned a RCT. None of the trials included in our study was from the mega journal PLOS One. We found 93 publications identified as registered reports and reporting results from RCTs. Most of these reports (92 out of 93) were published in the JMIR or its companion journals. Some of these papers had protocols published in the JMIR Research Protocols. According to JMIR policy, these protocol papers are peer reviewed and their acceptance “guarantees the subsequent acceptance of papers containing results from the protocol in any JMIR publications journal” (22). It is important to note that we also found registered reports for which the protocol was published in other journals, such as BMJ open, a journal that does not systematically implement peer reviews of protocols (23).

Unfortunately, we were not able to identify the dates of in-principle acceptance in the published articles. It would be relevant for this data to become standard in the reporting of registered reports, and also for publishers/editors to consider the interest of adding this to the article’s metadata. Rather than being typical registered reports, the publications tended to be RCTs with a published protocol, generally published or even formally accepted by peer review after inclusion of the first patient. However, protocols were generally submitted after the start of the study. This goes against the very principle of registered reports. Indeed, registered reports are not only tools that can deal with selective outcome reporting, they are also a means to enhance a given trial methodology, providing a good quality peer review. When a study is started, it is already too late to determine critical points regarding the methodology. Therefore, we could argue that a posteriori submission/acceptance of protocols is a major deviation from the original concept of registered reports. Furthermore, these protocols were often published in journals that were different from the journal publishing the final report. Of course, having a published protocol is surely a good thing for a RCT, since it should be associated with greater quality (24). For instance, the trials included in our survey tend to be larger than the prototypal RCT published in the field of biomedicine - as illustrated by a recent example on RCTs on interventions for mood, anxiety, and psychotic disorders (25), – most of which had sufficient power to detect effect sizes larger than 0.5. However, having a published protocol is not sufficient for it to become a registered report, especially when the protocol is published in a journal that is independent from the journal accepting the final report. In this case, despite having an IRRID, these final reports can hardly be considered as registered reports and at the very least they present major deviation. An even greater subject of concern is that some of the trials included were registered retrospectively on a RCT registry. Some of the publications with an IRRID reported on the results for secondary analyses. Concerning the reports of results from primary analyses, we identified frequent changes in the primary outcomes between the published protocol and the final publication. Few of these changes were explicitly discussed in the reports. It is often assumed, in classic papers, that outcome switching is a consequence of the pressure to publish positive results. Our results suggest that there are some other incentives at play, since researchers still switch outcomes, despite the guarantee they will be published.

### Strengths and weaknesses of this study

Our focus on RCTs makes sense because of the importance these studies have in evidence-based medicine. In addition, the proponents of registered reports have explicitly suggested that this format should be mandatory for RCTs because of the confirmatory nature of these trials (14). However, we should be careful to avoid any generalization of our results to other types of study, other journals and indeed to other fields. We found a highly selective sample of trials, derived from a specific journal group - JMIR focuses mainly on digital interventions - with its own policies. Impact and attention measures - such as citations and attention scores - suggest that the trials included were given a modest impact/media coverage.

Identifying registered reports is a difficult task. It is possible that our literature searches overlooked some references, as it has been previously noted that a substantial number of published final reports did not clearly identify themselves as registered reports (26). In contrast, the use of IRRID may have captured JMIR publications more easily. During the peer-review process, we also used terms such as “registered replication report*” and “preregistered report*” and found no additional reference.

One interesting perspective or “mise en abîme” would have been to publish this study as a registered report, but on this occasion, we chose not to, as it was more a descriptive survey, not based on hypothesis testing. To ensure transparency and reproducibility, the study was registered before data collection and all data, and all references included, are shared on the Open Science Framework.

### Strengths and weaknesses in relation to other studies

Previous studies on registered reports have highlighted a number of implementation issues and have suggested developing standards to ensure optimal implementation of this new publication format (14,26). These standards are definitely needed for registered reports on RCTs. Of course, there is a continuum of practices between protocol registration, protocol publication and indeed the registered report (14) and some of the practices at JMIR may be good for reproducibility purposes. Still, very particular attention should be paid to the use of the term registered report or to the assignment of a registered report identification number to these studies.

While registered reports are believed to strengthen the integrity of the publication landscape, it must be acknowledged that evidence of their real impact is still preliminary. One study has specifically attempted to evaluate the quality of registered reports in comparison to non-registered report papers in psychology and neuroscience, with positive results regarding the soundness of the registered report methodology and overall quality (27). Another study in the psychology literature shows a lower proportion of positive results in registered reports than in the literature overall, suggesting a lower level of publication bias with registered reports (28). Regarding the impact of registered reports, an ongoing study shows that they could be associated with more citations, but this data is preliminary and it could be related to the originality of the format rather than the content the reports that attracts these citations (29).

Future development will require meta-research to demonstrate the beneficial impact of this format. A collaboration involving journals and the Center for Open Science (COS) is underway to conduct a large, pragmatic, randomized controlled trial on registered reports compared to standard practice, to assess their impact on publication, research outcomes, and methodological quality (30).

### Meaning of the study: possible explanations and implications for clinicians and policy-makers

Overall, these results are compelling: not only is therapeutic research lagging behind with regard to the implementation of registered reports (26), but their implementation to date is far removed from the very idea of a registered report. With just a few exceptions, trialists interested in adopting this format cannot implement it because the journals - especially the leading general medical journals - do not provide this opportunity. Planning for registered reports is a first step towards a much-needed open science pathway for RCTs (6). The development of a model of this sort is not without risk for journals, and specific methodological developments relating to specific features of RCTs need to be considered. For instance, these trials are usually long and entail a risk of premature trial discontinuation (24). There is also a need to coordinate the various peer reviews that a trial receives in its life-cycle, e.g. before funding, by the IRB, or by health authorities, in order to make the process as efficient as possible. Reporting guidelines such as CONSORT (31) could also evolve to fit the needs of this new publication format.

### Unanswered questions and future research

Registered reports were developed as a means to improve research trustworthiness. We encourage key RCT funders, the main health authorities and major medical journals to join forces to develop piloting systems registered reports for RCTs. We cannot be satisfied with the current status quo for RCTs, where the concept of registered reports is almost inexistent, if not subverted. In addition, once registered reports are progressively implemented on a large scale, meta-research will be needed to evaluate how different journals and disciplines implement the format. The greater the number of adoptions of the format, the greater the risk of deviation from the general registered report policies.

## Supporting information

Web appendix 1

Web appendix 2

## Data Availability

All data produced are available online at https://osf.io/zf53p/

## WEB-APPENDIX

**Web appendix 1**: Validation of the procedure for registered report selection

**Web appendix 2**: Sources of extracted data

