## Supplementary figures and images for "Published registered reports are rare, limited to one journal group and inadequate for randomized controlled trials in the clinical field"

### Web appendix 1

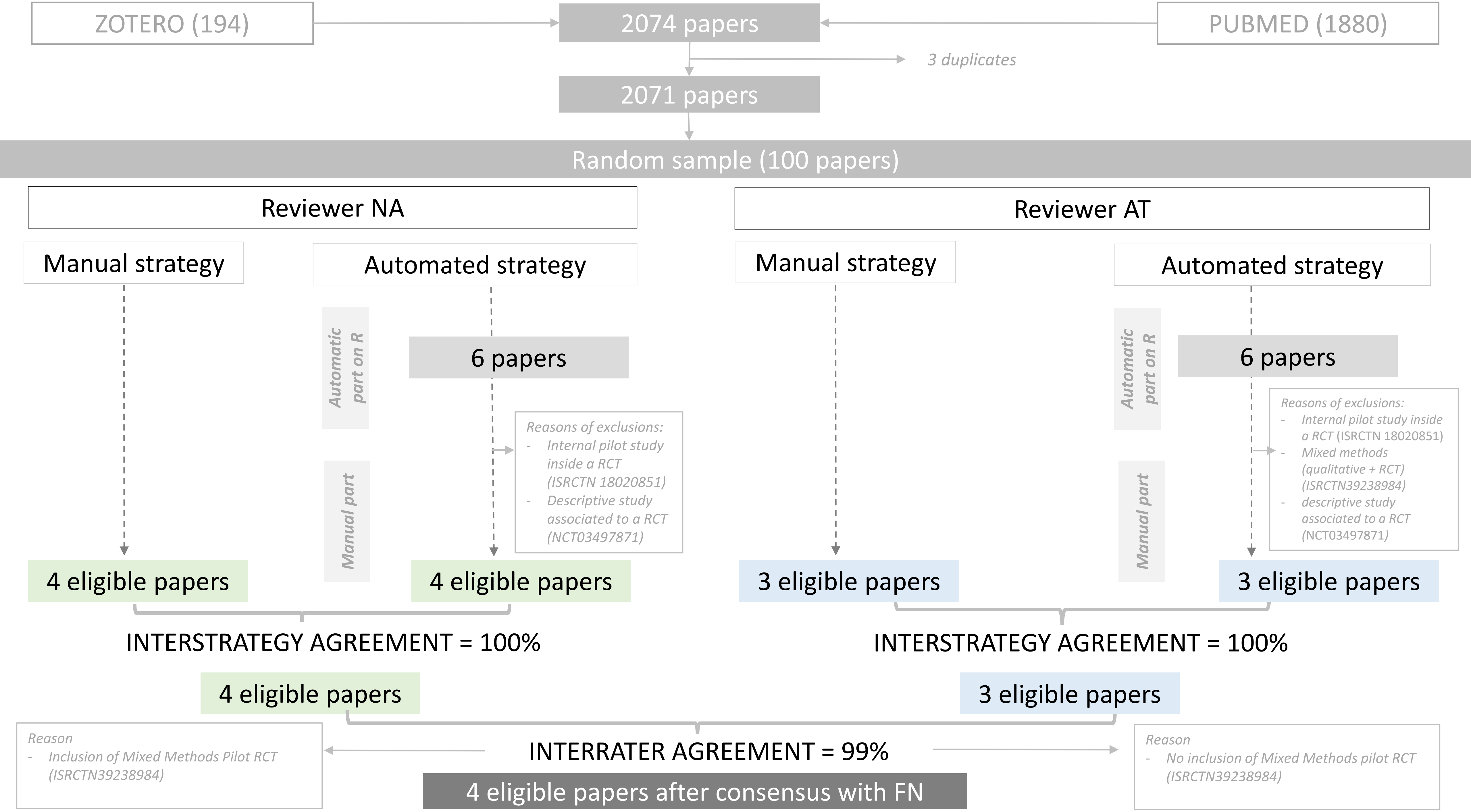
