## Supplementary material for "Published registered reports are rare, limited to one journal group and inadequate for randomized controlled trials in the clinical field": Web appendix 2

| **Extracted data** | **Site of extraction** |
| --- | --- |
| Date of study registration | Registry (ClinicalTrials.gov, ISRCTN registry, ANZCTR…etc) |
| Date of protocol submission | PubMed metadata |
| Date of protocol acceptance | PubMed metadata |
| Date of first patient inclusion | RR’s content |
| Date of protocol publication | PubMed metadata |
| Citations count | Web of Science |
| Altmetric Attention score | Altmetric bookmarklet |
| Rest of extracted data | PubMed metadata and RR’s content |
